# Before Birth, Beyond Childhood: Understanding the Influence of Prenatal Substance Exposure on Psychiatric Diagnoses

**DOI:** 10.64898/2026.05.27.26354273

**Authors:** Logan Caola, Evan Dastin-Van Rijn, Shelby Anderson, Erich Kummerfeld, Christi Sullivan, Stefanie Simpson, Aarav Kalkar, Ritwick Banerjee, Audrey Houghton, Mark Fiecas, Anita C Randolph

## Abstract

**Background:** Prenatal substance exposure (PSE) occurs when an individual is exposed to substances *in utero.* PSEs may have lasting effects on mental health. We tested whether PSEs show threshold, cumulative, or individual substance associations with childhood psychiatric diagnoses.

**Methods:** Clinical variables (demographics, ICD-9/10 diagnoses, PSE history) were extracted from electronic health records from the University of Minnesota Adoption Medicine Clinic. PSEs were identified from caregiver and child-protective-services narratives and/or toxicology (cord tissue/blood, meconium). For each ICD-9/10 diagnostic category, we fit logistic regression models comparing (1) exposure thresholds (0, 1, 2, 3, ≥4 exposures), (2) a cumulative exposure count, and (3) individual substances to estimate marginal odds ratios (ORs) with 95% Confidence Intervals (CIs).

**Results:** Psychiatric diagnoses increased with the number of PSEs. Relative to no exposure, odds of an Anxiety Disorder rose from OR 1.47 (95% CI 1.16–1.87) with one exposure to OR 2.03 (1.64–2.52) with ≥4 exposures. Higher cumulative exposure scores were associated with Anxiety Disorders (OR 1.28, 1.18–1.38), Behavioral and Emotional Disorders (OR 1.42, 1.31–1.54), Substance Use Disorders (OR 1.52, 1.29–1.79), and Mood Disorders (OR 1.16, 1.04–1.30). Alcohol, tobacco, and marijuana exposures were associated with increased odds of at least one psychiatric diagnosis, and each substance showed at least one significant diagnostic cluster when modeled independently.

**Conclusion:** Increasing numbers of PSEs were associated with higher odds of psychiatric diagnoses, with patterns varying by substance and outcome. These findings motivate research on exposure timing and combinations to support earlier identification and intervention for at-risk children.

## Background

Substance use during pregnancy is relatively common, with nearly one in five pregnant people having used nicotine, alcohol, or illicit substances such as heroin and methamphetamine in the past year, and one in ten meeting Substance Use Disorder (SUD) criteria in the past year.^1^ From 2000-2022, 380,000 children were born exposed to illicit substances, 500,000 exposed to alcohol, and over one million exposed to nicotine.^2^ In the adoption and foster care population there is a higher prevalence of prenatal substance exposure (PSE) compared to the general population, with an estimated 30% of newborns diagnosed with PSE at birth entering foster care at some point in their life.^3,4^ New evidence suggests that children with PSEs have increased risks for both short- and long-term adverse mental and physical health outcomes.^5^ Specifically, PSE is associated with an increased prevalence of diagnoses such as ADHD, intellectual disabilities, Mood Disorders, and behavioral problems.^6–11^ Studying PSE is methodologically challenging due to the heterogeneous exposure profiles and varying availability of objective measures of use like toxicology screening and self-reports, further complicating the interpretation and generalizability of results.^12,13^

We propose utilizing three adverse childhood experiences (ACEs) frameworks to explore the effects of PSE on later psychiatric diagnoses: the threshold risk model, cumulative risk model, and individual risk model. The threshold risk model states that as the number of ACEs increases, so too does the burden of adverse outcomes, with studies showing a significant association between higher ACE scores and earlier initiation of illicit and legal substances, illicit drug use problems, and development of addiction to illicit substances.^14,15^ The cumulative risk score model sums exposure to individual risk factors to generate a total risk score, demonstrated by a significant positive association between cumulative ACEs and substance use.^16–19^ Finally, the individual risk model examines ACEs separately as unique indicators of risk for psychopathology and other psychiatric diagnoses, such as prenatal cocaine exposure and its association with cognitive impairments, lower IQ, and motor skill deficits.^17^ Historically, these frameworks use ACEs and psychopathology for analysis; to our knowledge, we are the first study to explore if they can be used to examine long-term risk of psychiatric diagnoses following PSE.

We aim to leverage data from our Building Understanding of Transitions & Trauma in Early Formative Life Years (BUTTERFLY) and Clinical Outcomes & Medical Profiles of Children in Adoption & Statecare Settings (COMPASS) datasets (electronic health records; EHRs) databases from the University of Minnesota’s Adoption Medicine Clinic (UMN-AMC) to contribute a deeper understanding of the risks that PSE confers for psychiatric diagnoses.^4^ We hypothesize that the number of psychiatric diagnoses will increase as the number of PSEs increases.

## Methods

### Study Participants

The study population was patients for whom PSE and health data were available in EHRs from the UMN-AMC. The UMN-AMC serves children who experience higher rates of early-life adversities than the general population, including children currently in the foster care system, those recently adopted domestically and internationally, and adopted children with severe milestone delays and mental health disorders.

### Design and data extraction

Using a standardized operating procedure, five trained Research Professionals extracted data from 3,685 patients ages 0-21 years old who completed their first Comprehensive Child Wellness Assessments (CCWA) from 2014 through 2024. Measures extracted include demographic, PSE history, and International Classification of Diseases, 9th and 10th edition (ICD-9 / ICD-10) codes for psychiatric disorders. ICD-9/10 F codes were collected directly from patients’ EHRs via the UMN’s Best Practice Informatics Core (BPIC) staff, who compiled a list of all diagnoses of both medical and psychiatric for each individual participant from their entire medical history. Diagnostic outcomes can be found in Table 1. Eligible patients were excluded from analyses if patients’ EHR data were inaccessible or incomplete (eg, for patients who either did not attend or schedule their follow up appointment(s) or new to the clinic and had no historic clinical data from 2014-2024). This resulted in a final analytical sample of n = 2720.

**Table 1.**
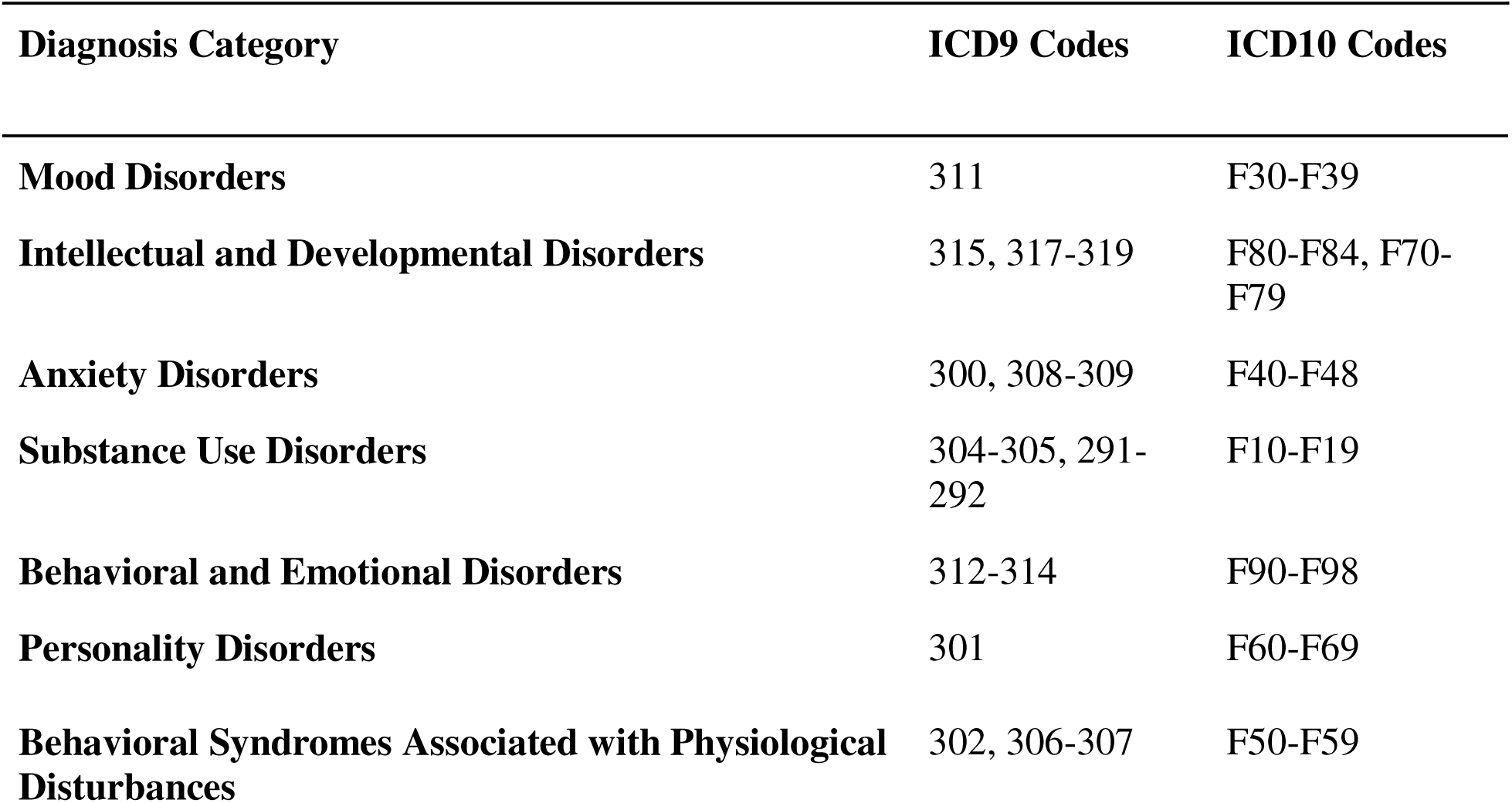
Mapping of diagnostic categories to ranges of ICD codes.

## Measures

### Demographic data

Data was collected for patient gender (sex), age at initial visit, and race. Ethnicity data was recorded by practices using 2020 US census categories but were excluded in analyses due to missing data (>50% of total population).

### Prenatal substance use exposure

PSEs were documented as confirmed (yes/no), suspected, or testing not available before or during the first CCWA at the UMN-AMC visit. Caregivers, social workers, and/or Child Protective Services case workers provided history of PSE in narrative form and/or provided cord tissue, cord blood, and/or meconium toxicology screening results. Although 10 exposure data variables were collected for numeric analysis, only six were used for individual exposure analysis: alcohol, tobacco, marijuana, methamphetamine, opioids, and cocaine. The opioids category was combined, encompassing illicit opioids (heroin, fentanyl, prescription pain pills) and medications for opioid use disorder (Suboxone, Methadone, Sublocade).

### Statistical Analysis

Primary analysis was carried out on cross-sectional data extracted from the UMN-AMC EHRs. All available diagnosis data was grouped according to the initial three characters of the ICD code. Codes were further grouped into larger diagnostic categories. A full mapping of ICD codes to diagnosis labels is provided in Table 1. The ten PSE categories collected were further grouped into alcohol, tobacco, marijuana, methamphetamine, opioids, and cocaine. Each diagnostic category was then used as the outcome variable for a set of logistic regressions, with a ‘1’ indicating the child had at some point in their life received a diagnosis, confirmed PSE, or were suspected of PSE and a ‘0’ indicating that the child did not receive the diagnosis, nor PSE over the period measured in the database. For each diagnosis, we fit three types of models. The first model tested relations between the threshold risk model and diagnoses, with each count (1, 2, 3, 4 or more) compared against a reference group who had 0 exposures. A second model estimated the associations between a cumulative PSE score and diagnoses for all possible substance exposures from the UMN-AMC data. Weighted score values were capped to a range from 0-10 to prevent rare extreme values from driving the coefficient. Less than 1% of the children had more than ten prenatal exposures. In the third model, we estimated univariate associations between the six most common types of PSEs and diagnoses. ‘Sex’ and ‘Age at Initial Visit’ were included as additional predictors in each model to handle confounds. P-values were adjusted for a false discovery rate of 0.05 using the Benjamini-Hochberg procedure to handle multiple comparisons. Coefficients and 95% confidence intervals (CI) from the logistic regression were transformed into odds ratios (OR) when presenting the data. For the weighted score, odds ratios are reported per one standard deviation increase to facilitate comparison with categorical predictors.

## Results

Results from descriptive measures are presented in Table 2. Among the psychiatric diagnoses assessed in the study, the most prevalent were Anxiety Disorders (60.3%) and Intellectual and Developmental Disorders (47.9%). The least prevalent were Personality Disorders (3.9%) and Substance Use Disorders (4.9%). PSEs to various substances were highly prevalent in the dataset, with 71.9% of the children having an exposure to at least one substance, 56.1% having at least two, 39.5% having at least three, and 25.2% having four or more. Among exposures, alcohol (53.7%) and tobacco (32.1%) were the most prevalent, while cocaine (11.8%) and opioids (14.7%) were the least prevalent.

**Table 2.** Description of study measures (N=2720)

| Variable | count (%) or mean [SD] |
| --- | --- |
| **Demographics** |  |
| Age at Initial Visit (range 0-21) | 7.2 [4.4] |
| Female | 1290 (47.4) |
| **Mental Health Diagnoses** |  |
| **Mood Disorders** | 421 (15.5) |
| **Intellectual and Developmental Disorders** | 1303 (47.9) |
| **Anxiety Disorders** | 1640 (60.3) |
| **Substance Use Disorders** | 133 (4.9) |
| **Behavioral and Emotional Disorders** | 1266 (46.5) |
| **Personality Disorders** | 105 (3.9) |
| **Behavioral Syndromes Associated with Physiological Disturbances** | 218 (8.0) |
| **Prenatal exposures** |  |
| **Alcohol** | 1460 (53.7) |
| **Tobacco** | 872 (32.1) |
| **Marijuana** | 853 (31.4) |
| **Methamphetamine** | 715 (26.3) |
| **Opioids** | 399 (14.7) |
| **Cocaine** | 320 (11.8) |
| **Cumulative exposure score (range 0-10)** | 2.3 [2.2] |

*Note.* ACEs = adverse childhood experiences. SD = Standard deviation.

Table 3 presents the results from logistic regressions testing relations between threshold and cumulative PSE count and psychiatric diagnoses. Modeling showed increasing numbers of exposures were generally associated with increased odds of poor psychiatric diagnoses. To illustrate, compared to children with no PSEs, the odds of receiving an Anxiety Disorder Diagnosis was 1.47 (CI = 1.16-1.87) with one exposure, 1.84 (CI = 1.45-2.34) with two exposures, 1.90 (CI = 1.47-2.44) with three exposures, and 2.03 (CI = 1.64-2.52) with four or more exposures. A second set of regression models also indicated that total exposure score was associated with Mood Disorders (OR = 1.16, CI = 1.04-1.30), Anxiety Disorders (OR = 1.28, CI = 1.18-1.38), Substance Use Disorders (OR = 1.52, CI = 1.29-1.79), Behavioral and Emotional Disorders (OR = 1.42, CI = 1.31-1.54), Personality Disorders (OR = 1.28, CI = 1.07-1.54), and Behavioral Syndromes Associated with Physiological Disturbances (OR = 1.27, CI = 1.12-1.44).

**Table 3.**
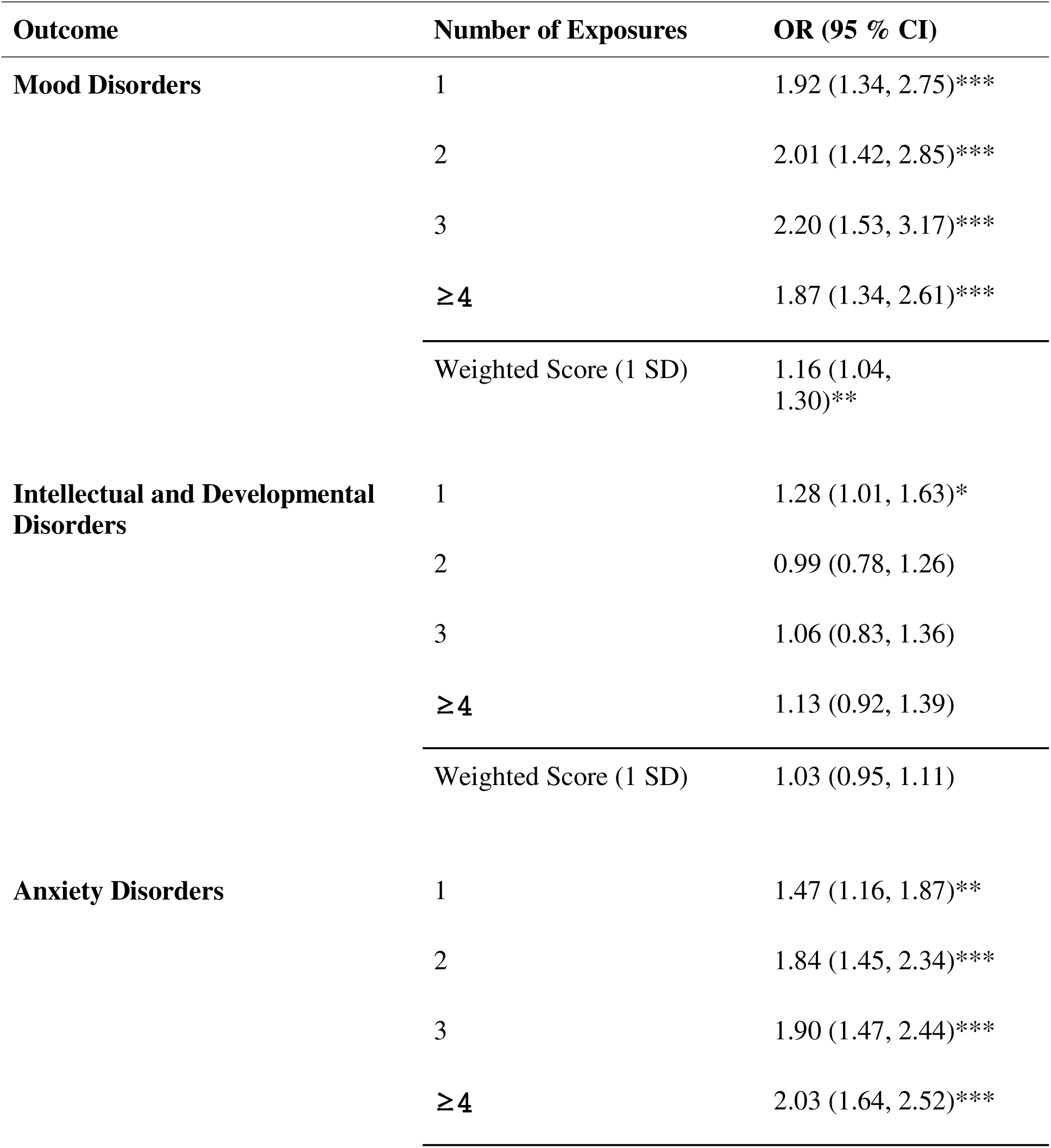
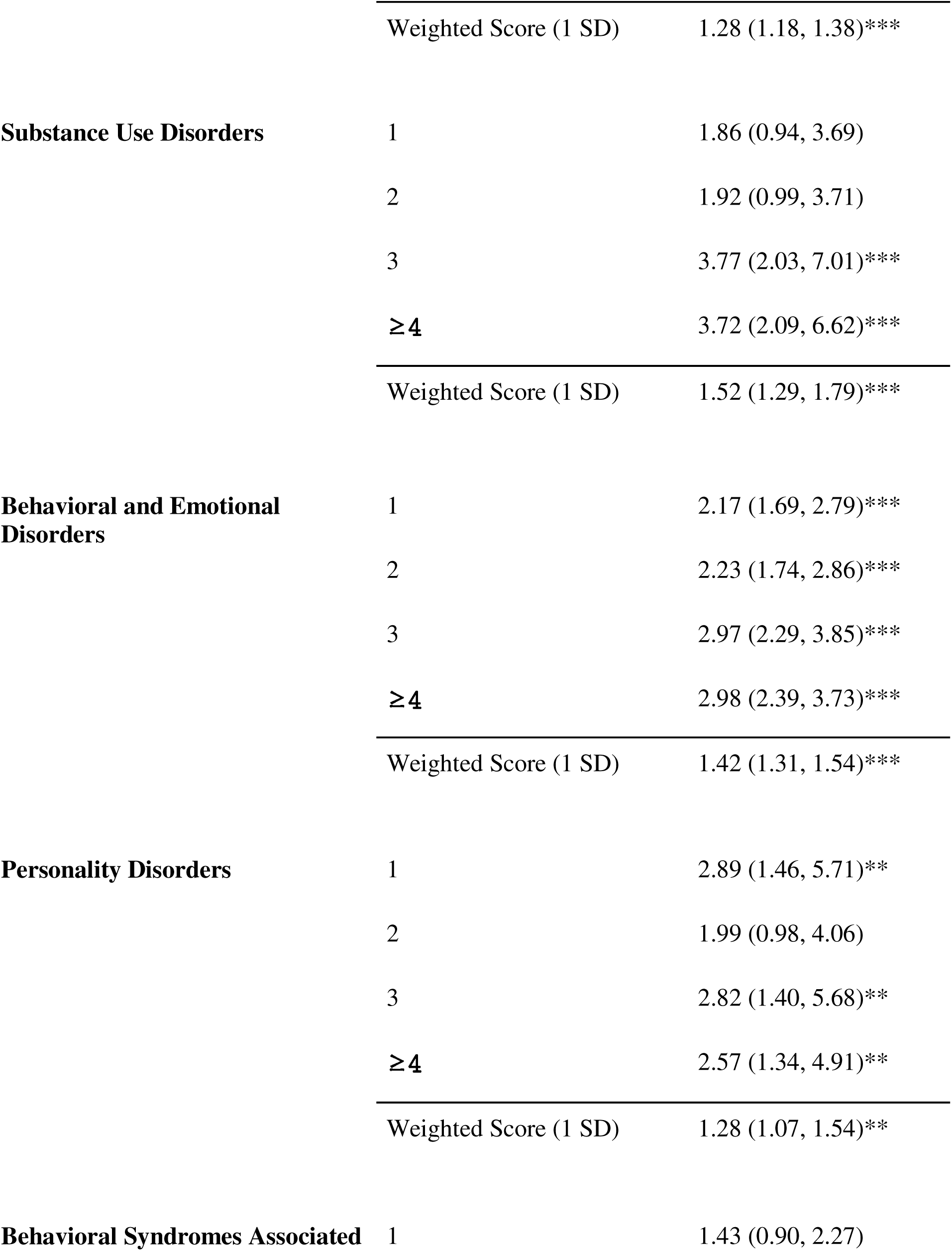
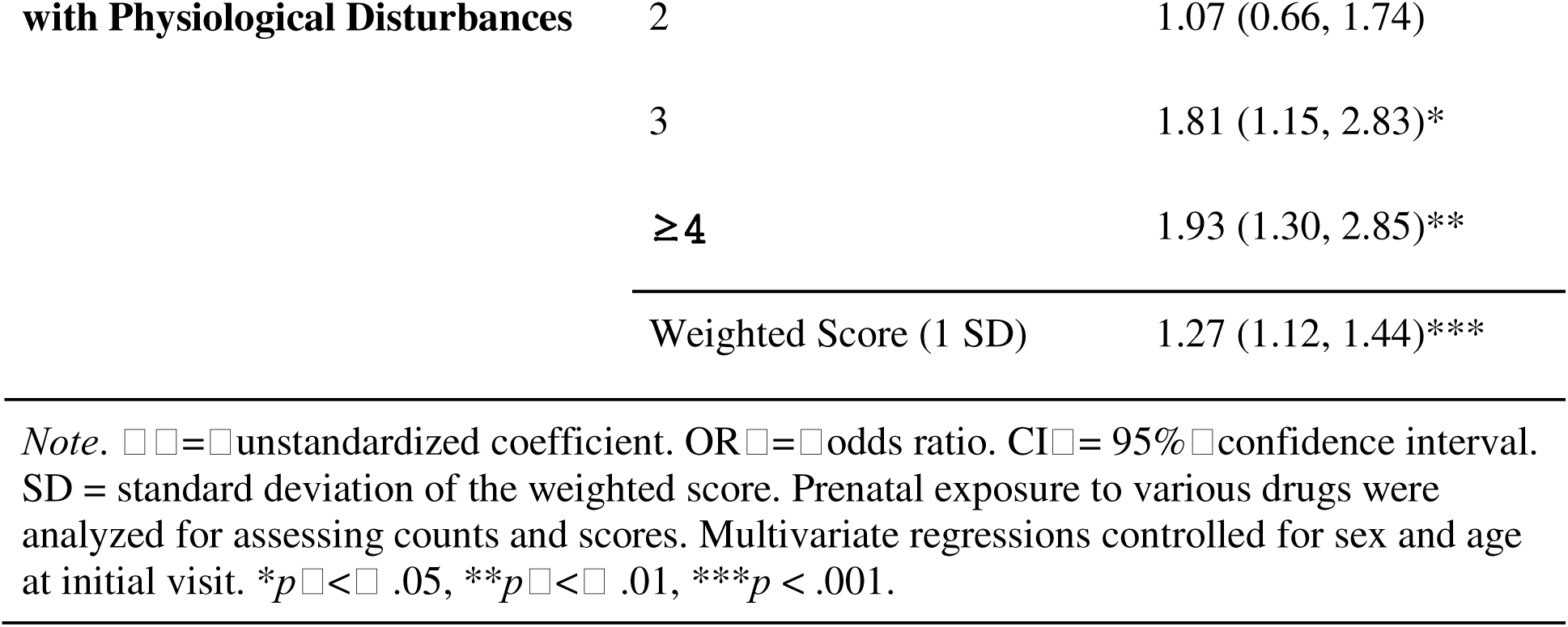
Associations between prenatal exposure counts and psychiatric diagnoses. Threshold Risk Model = 1, 2, 3, ≥4 exposures; Cumulative Score Risk Model = Weighted Score.

Results in Table 4 are from a series of logistic regressions associating individual types of PSEs with psychiatric diagnoses. Across diagnoses, individual exposures had substantial and significant associations with increased risk particularly alcohol, tobacco, and marijuana. Effect sizes and CIs for all exposure type coefficients are visualized in Figure 1B.

**Figure 1:**
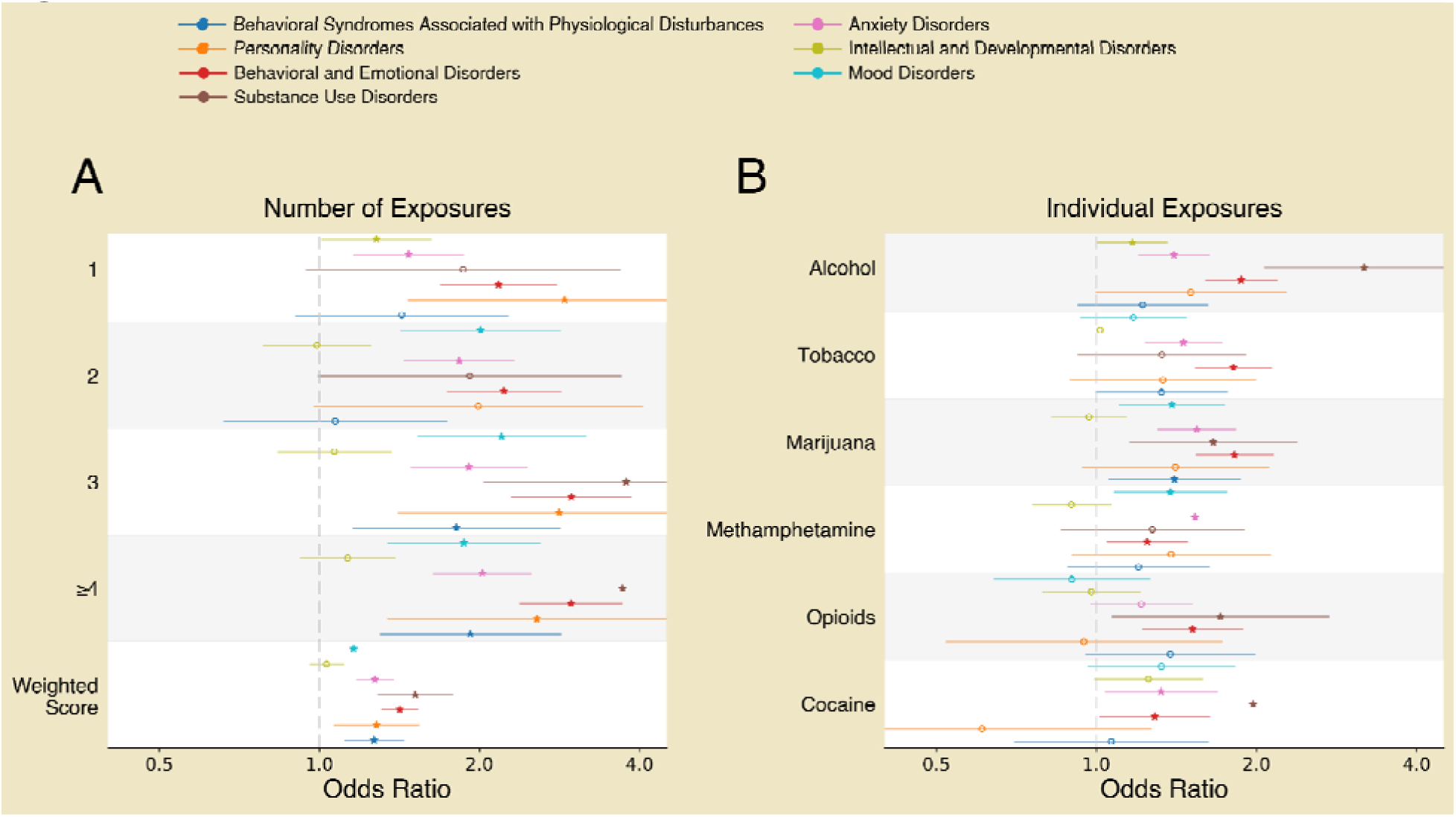
Forest plot of marginal effects of PSE counts and types on psychiatric and neurological indications. Each panel shows estimated effects and 95% CIs for prenatal exposure counts and types across 7 diagnostic outcomes. Points indicate mean parameter estimates for odds ratios, and horizontal lines denote corresponding CIs. Significant predictors are indicated by star markers. Outcomes for each exposure feature are grouped by row. Panels correspond to different groupings of exposure features (**A** - Number of prenatal exposures, **B** - Exposure type). For the exposure score, the odds ratio corresponds to a one standard deviation increase. Individuals were assigned “1” if they had at least 1 confirmed or suspected exposure within the exposure type group and a “0” if they had no exposure.

**Table 4.** Associations between types of prenatal exposures and neuropsychiatric diagnoses. Individual Substance Risk Model.

| Outcome | Individual Exposures | OR (95 % CI) |
| --- | --- | --- |
| **Mood Disorders** | Alcohol | 1.40 (1.12, 1.76)** |
|  | Tobacco | 1.17 (0.93, 1.48) |
|  | Marijuana | 1.39 (1.10, 1.75)** |
|  | Methamphetamine | 1.38 (1.08, 1.77)* |
|  | Opioids | 0.90 (0.64, 1.27) |
|  | Cocaine | 1.33 (0.96, 1.83) |
| **Intellectual and Developmental Disorders** | Alcohol | 1.17 (1.00, 1.36)* |
|  | Tobacco | 1.01 (0.86, 1.19) |
|  | Marijuana | 0.97 (0.82, 1.14) |
|  | Methamphetamine | 0.89 (0.75, 1.06) |
|  | Opioids | 0.97 (0.78, 1.21) |
|  | Cocaine | 1.25 (0.98, 1.57) |
| **Anxiety Disorders** | Alcohol | 1.40 (1.20, 1.64)*** |
|  | Tobacco | 1.46 (1.24, 1.73)*** |
|  | Marijuana | 1.54 (1.30, 1.83)*** |
|  | Methamphetamine | 1.54 (1.28, 1.84)*** |
|  | Opioids | 1.22 (0.98, 1.53) |
|  | Cocaine | 1.34 (1.04, 1.71)* |
| **Substance Use Disorders** | Alcohol | 3.19 (2.07, 4.92)*** |
|  | Tobacco | 1.33 (0.92, 1.92) |
|  | Marijuana | 1.66 (1.15, 2.39)** |
|  | Methamphetamine | 1.28 (0.86, 1.90) |
|  | Opioids | 1.71 (1.07, 2.75)* |
|  | Cocaine | 1.98 (1.25, 3.12)** |
| **Behavioral and Emotional Disorders** | Alcohol | 1.87 (1.60, 2.20)*** |
|  | Tobacco | 1.81 (1.54, 2.15)*** |
|  | Marijuana | 1.82 (1.54, 2.15)*** |
|  | Methamphetamine | 1.25 (1.05, 1.49)* |
|  | Opioids | 1.52 (1.22, 1.90)*** |
|  | Cocaine | 1.30 (1.02, 1.65)* |
| **Personality Disorders** | Alcohol | 1.51 (0.99, 2.28) |
|  | Tobacco | 1.34 (0.89, 2.00) |
|  | Marijuana | 1.41 (0.94, 2.11) |
|  | Methamphetamine | 1.38 (0.90, 2.13) |
|  | Opioids | 0.95 (0.52, 1.73) |
|  | Cocaine | 0.61 (0.29, 1.27) |
| **Behavioral Syndromes Associated with Physiological Disturbances** | Alcohol | 1.22 (0.92, 1.62) |
|  | Tobacco | 1.33 (1.00, 1.77) |
|  | Marijuana | 1.40 (1.05, 1.87)* |
|  | Methamphetamine | 1.20 (0.88, 1.64) |
|  | Opioids | 1.38 (0.96, 2.00) |
|  | Cocaine | 1.07 (0.70, 1.63) |

*Note.* OR = odds ratio. CI = 95% confidence interval. Each cell shows the odds ratio and confidence interval for the corresponding coefficient. Multivariate regressions controlled for sex and age at initial visit. \* $p < .05$ , \*\* $p < .01$ , \*\*\* $p < .001$ .

Across the sample, there was substantial polysubstance exposure, and greater cumulative exposure was associated with progressively increased odds of adverse psychiatric outcomes across diagnoses. The analysis of individual substances indicated that alcohol, tobacco, and marijuana accounted for many of these associations and were the most consistently linked to risk. Together, the tables and figures highlight consistent predictors across diagnostic outcomes and reveal diagnosis-specific patterns in effect direction and magnitude.

## Discussion

Utilizing three ACE frameworks (threshold, cumulative score, and individual risk models), our study aimed to explore the association between PSE and their influence on later psychiatric diagnoses. This is, to our knowledge, the first time these frameworks have been used to study PSE’s effects on psychiatric outcomes. Consistent with our hypothesis, we found that as the number of PSEs increased, a significant increase in psychiatric diagnoses (i.e., Mood, Anxiety, Substance Use, Behavioral and Emotional, Personality, and Behavioral Syndromes Associated with Physiological Disturbances) was observed. Even for psychiatric diagnoses (i.e., Intellectual and Developmental Disorders) with non-significant findings, a trend of increased association was observed where more exposures increased the odds of a mental health diagnosis. Although our results corroborate previous findings, this study pushes the field forward because our database allowed us to investigate the impacts of PSEs on psychiatric diagnoses rather than symptoms, providing evidence that adverse exposures are associated with specific diagnoses later in life.

### Threshold Risk Model

Our data supports the threshold risk model (see Table 3, Figure 1A), which proposes that any increase in ACEs and PSEs leads to negative psychiatric diagnoses. As the number of PSEs for subjects increased, so did the risk for Mood Disorders, Anxiety Disorders, Substance Use Disorders, Behavioral and Emotional Disorders, Personality Disorders, and Behavioral Syndromes Associated with Physiological Disturbances, demonstrating a dose-response relationship between PSE and these psychiatric diagnoses. Due to the presence of any PSE, the risk of psychiatric disorders increased with increased exposure to PSEs. These results provide evidence that adverse events that occur during the prenatal development environment can impact the child’s development.

### Cumulative Score Risk Model

The cumulative risk model, which postulates that exposure to higher numbers of PSE leads to increased prevalence of negative outcomes, was also supported by our study (see Table 3, Figure 1A). Several large data studies show a cumulative effect from PSEs and psychiatric diagnoses, such as the ABCD Study.^18^ Our data shows that a high cumulative score is associated with Mood Disorders, Anxiety Disorders, Substance Use Disorders, Behavioral and Emotional Disorders, Personality Disorders, and Behavioral Syndromes Associated with Physiological Disturbances. This means that diagnoses within each listed cluster become more prevalent as the number of PSEs increases. These results support the idea that one exposure is enough to impact health outcomes, but adding more PSEs further increases the risk for significant health outcomes.

### Individual Substance Risk Model

The individual substance risk model speculates that individual PSE are unique indicators of risk for negative outcomes. Our data demonstrates that exposure to specific substances can lead to unique mental health profiles, which aligns with previous research (see Table 4, Figure 1B). Prenatal alcohol exposure has been previously shown to cause externalizing symptoms and behavioral problems, and our results shows it is significantly associated with Mood Disorders, Intellectual and Developmental Disorders, Anxiety Disorders, Substance Use Disorders, and Behavioral and Emotional Disorders.^20^ Prenatal nicotine exposure in our data is linked to Anxiety and Behavioral and Emotional Disorders, consistent with evidence connecting nicotine to both internalizing and externalizing symptoms and broad psychiatric morbidity.^21,22^ Similarly, our data also find significant associations with marijuana exposure with Mood Disorders, Anxiety Disorders, Substance Use Disorders, Behavioral and Emotional Disorders, and Behavioral Syndromes Associated with Physiological Disturbances, though links to prenatal marijuana exposure and higher rates of psychotic-like experiences and social problems were not examined in this study.^6,10^ A study by Roffman et al., found that increased frequency of prenatal exposure to tobacco, alcohol, and marijuana led to a significant increase in internalizing symptoms, such as Anxiety and Depression, and externalizing symptoms (aggression, attention problems, behavioral issues).^24^

Our data also suggests that prenatal opioid exposure is associated with Substance Use Disorders and Behavioral and Emotional Disorders, while methamphetamine is associated with Mood Disorders, Anxiety Disorders, and Behavioral and Emotional Disorders. This corroborates findings from prior studies showing that the use of illicit substances in pregnancy, such as opioids, cocaine, and methamphetamine, were found to increase externalizing behaviors such as attention and behavioral problems.^9,23^ Methamphetamine in particular was associated with increased emotional reactivity, attention deficits, hyperactivity, and anxious or depressed problems.^9^ Likewise, exposure to cocaine in pregnancy (another stimulant) is associated with higher odds of increasing Anxiety Disorders, Substance Use Disorders, and Behavioral and Emotional Disorders in our data. This correlates with previous research that found methamphetamine and cocaine affect emotional reactivity and increases Anxiety (F40) symptoms and disposition.^7,9^

In total, our data supports that there are unique profiles of mental health risks based on the PSE. It is unclear if these profiles remain consistent across populations. For example, our results found PSE of marijuana to be associated with Mood Disorders and Anxiety Disorders. One meta analysis found that prenatal marijuana exposures in infants was associated with psychiatric diagnoses, externalizing problems, and attention problems which is similar to our findings, but another found that there were no associations between marijuana and anxiety or depression which is contradictory to our findings.^23,25^ Additionally, only the PSE of alcohol was significantly associated with Intellectual and Developmental Disorders despite a cluster of other studies finding that exposures to alcohol, tobacco, marijuana, cocaine, and opioids are related to Intellectual and Developmental Disorders.^8,26–31^ Further investigation and comparisons of study population, exposure types, and analytic approach is warranted to further understand if individual PSEs have unique profiles or if there are additional factors that influence which mental health outcomes are most associated with specific PSEs.

## Conclusion

The key takeaway of this study is the significant relationship between PSEs and elevated risk for various psychiatric diagnoses. Analyses identified key relationships between specific substances and specific clusters of psychiatric outcomes, creating a potential area of research and clinical care focused on how specific substances lead to specific diagnoses. Our findings justified increased intervention points across pregnancy, infancy, and early childhood.

## Limitations

The majority of the data collected from our database comes from first-person sources, such as birth toxicology or bio-parent self report. However, a mix of secondary sources (friends of the patient, foster family, relatives) also confirmed PSE, often reporting suspected exposure. Thus, secondary sources could result in underreporting of exposure counts. Another limitation of this data is that all exposures are considered on a categorical scale: exposed, suspected, or not-exposed. In reality, there are more nuances to PSEs, including maternal intensity of usage, timing of usage in relation to pregnancy, and polysubstance use. Consequently, mono-exposure data was limited, which may impact the diagnoses clusters generated for specific PSEs. Finally, many families who seek evaluations from the UMN-AMC do so to get access to resources like educational and disability services. The clinic is open to all adoption/foster care children in the Midwest, but only those already experiencing behavioral or health difficulties may be more likely to follow through with appointments. This population may skew the data to overrepresent negative psychiatric outcomes, suggesting that further research with a full age matched control population is needed to improve generalizability.

## Future Directions

One area that warrants future research is examining synergistic effects of different PSEs on psychiatric diagnoses. For example, children exposed to both marijuana and opioids show a greater prevalence of ADHD than expected from either exposure individually.^32^ Identifying which specific combinations of PSEs could lead to specific types of psychiatric diagnoses, improving targeted preventative care and coordinated intervention. Additionally, researching how timing and duration of PSEs relate to psychiatric diagnoses would provide further context into the type of preventative care and interventions needed.^33^

Examining moderating factors, like demographics, caregiving environment, and personal experiences is needed as foster care has been shown to exacerbate psychiatric diagnoses due to caregiver disruption and other adverse childhood experiences.^34^ Knowing which factors of upbringing can exacerbate future psychiatric diagnoses would also provide a groundwork necessary for successful preventative care.

Policy reform supporting prevention frameworks and early interventions for those with PSE is essential. Introducing additional check-ins for mothers experiencing substance use, and providing motivation to allow for the honest and accurate reporting of frequency and intensity of substances without repercussions, could be used to create the necessary baseline for a child born with PSE. Non-punitive reporting environments and additional early life screening procedures may improve identification of PSE-related diagnostic profiles and earlier access to medical and educational interventions.

## Data Availability

All data is available online at https://conservancy.umn.edu/browse/author?value=Randolph,%20Anita&bbm.return=1

## Declarations

## Ethics approval and consent to participate

This study was conducted under the approval and oversight of the Institutional Review Board (IRB STUDY00019280) at UMN. All procedures were in compliance with federal regulations, institutional policies, and ethical guidelines for human research. No informed consent was required for this retrospective chart review.

## Consent for publication

Not applicable

## Availability of data and materials

De-identified participant data—including baseline demographics, prenatal exposures, psychiatric diagnoses, and more, are available for analysis in the <u>Data Repository for the University of Minnesota (DRUM)</u>. The two de-identified datasets used in this study Building Understanding of Transitions & Trauma in Early Formative Life Years (BUTTERFLY) and Clinical Outcomes & Medical Profiles of Children in Adoption & Statecare Settings (COMPASS) can be found at these respective links: https://hdl.handle.net/11299/279102; https://hdl.handle.net/11299/279103

This data is protected by an attribution noncommercial sharealike 4.0 international license. You must give appropriate credit, provide a link to the license, and indicate if changes were made. You may not in any way that suggests the licensor endorses you or your use. You may not use the material for commercial purposes. If you remix, transform, or build upon the material, you must distribute your contributions under the same license as the original. You may not apply legal terms or technological measures that legally restrict others from doing anything the license permits.

## Competing interests

MF reports grants from National Institutes of Health, Food and Drug Administration, and National Science Foundation. All other authors declare no competing interests.

## Funding

This research was supported by the University of Minnesota’s Data Science Initiative, Department of Pediatrics, Masonic Institute for the Developing Brain, Children’s Discovery Fund, and the National Institutes of Health’s National Center for Advancing Translational Sciences, grant UM1TR004405. This funding had no involvement on the study design, analysis, interpretation, writing, or submission of this manuscript.

## Authors’ contributions

AR and MF conceived the project. AR secured funding. AR, LC, and SA supervised data capture, entered data manually, and validated data of enrolled patients. LC and SA provided access to the dataset. ED-VR, AK, RB, and AR curated the data. EK, MF, and AR supervised initial data entry and analysis. ED-VR and AH supervised data analysis and led data clustering and analysis with outcomes, and data visualization. ED-VR and AH analysed data, including data modelling and statistical analysis. All authors had access to the raw data. ED-VR and LC wrote the initial draft of the manuscript. LC, SA, CS, SS, and AR wrote the final manuscript. All the authors reviewed the final version of the manuscript and had full access to all data reported in the study. ED-VR, AH, AK, and RB verified the data.

## Acknowledgements

We thank all the staff of the University of Minnesota, Adoption Medicine Clinic. We would also like to thank the individuals that helped lay the initial methodological framework for this data, as well as helped with data entry and quality control: Taylor Rawstern, Jada Loleng, Jonathan Lehman, Jade Roghair

## List of Abbreviations

PSE: Prenatal substance exposure
SUD: Substance Use Disorder
ACEs: Adverse Childhood Experiences
EHRs: Electronic Health Records
UMN-AMC: University of Minnesota’s Adoption Medicine Clinic
CCWA: Comprehensive Child Wellness Assessments
ICD-9 / ICD-10: International Classification of Diseases, 9th and 10th edition
BPIC: Best Practice Informatics Core
OR: Odds Ratios
CI: Confidence Intervals

## Notes

### Competing Interest Statement

The authors have declared no competing interest.

### Author Declarations

The IRB of the University of Minnesota Twin Cities gave ethical approval for this work

### Summary of Updates

The personal cell phone number was not removed prior to submission

